# Prevalence and Correlates of Malnutrition and Anaemia among Young Children in Rural Northeastern Ghana: A Community-Based Cross-Sectional Study

**DOI:** 10.1101/2025.11.17.25340375

**Authors:** Vincent Adocta Awuuh, Charles Apprey, Reginald Adjetey Annan

## Abstract

**Introduction:** Undernutrition remains a major public health concern in low- and middle-income countries, including Ghana, with generational consequences. This study examined the prevalence and place-based correlates of malnutrition and anaemia among children under five years old in rural Northeastern Ghana, aiming to develop effective interventions.

**Methodology:** A community-based cross-sectional study was conducted in 10 rural communities in the East Mamprusi Municipality in June/July 2023. Anthropometric and haemoglobin measurements were taken from children under five, alongside socio-demographic, maternal health, and feeding practice data. Descriptive and logistic regression were used to identify the prevalence and determinants of child nutrition indicators.

**Results:** The prevalence of malnutrition was 34.3% for stunting, 31.9% for wasting, 27.8% for underweight, and 55.2% for anaemia. Key associated factors of underweight included a lack of home healthcare provider support [AOR = 21.44, 95% CI: 3.39–135.78, p = 0.001] and non-exclusive breastfeeding [AOR = 9.53, 95% CI: 1.71–53.06, p = 0.01]. Wasting was associated with the absence of age-appropriate complementary feeding [AOR = 11.23, 95% CI: 2.35–53.74, p = 0.002] and failure to prevent iodine deficiency [AOR = 8.37, 95% CI: 1.93–36.24, p = 0.004]. Stunting was linked to lower household food expenditure [AOR = 0.97, 95% CI: 0.97–0.98, p < 0.001] and limited caregiver knowledge of exclusive breastfeeding [AOR = 0.38, 95% CI: 0.16–0.91, p = 0.03]. Anaemia was more likely in children whose mothers did not implement anaemia-preventive practices [AOR = 6.41, 95% CI: 1.31–31.32, p = 0.022], while protective factors included maternal satisfaction with health services [AOR = 0.13, 95% CI: 0.03–0.50, p = 0.003] and exclusive breastfeeding [AOR = 0.31, 95% CI: 0.10–0.93, p = 0.036].

**Conclusion:** Child malnutrition and anaemia remain unacceptably high in rural Northeastern Ghana. Co-designed, community-specific and culturally relevant strategies are urgently needed to address these challenges sustainably.

## Introduction

Undernutrition, manifesting as anaemia, underweight, wasting, or stunting [1; 2], remains a major global public health problem despite progress over the past decades [3]. It contributes to 45% of child mortality worldwide, impairs physical and cognitive development, and leads to productivity losses of up to 16% of gross domestic product in developing countries [4;1].

Globally, approximately one in four children under five is stunted, with Africa and Asia bear the highest burden of malnutrition among women and children: wasting (Asia [70%]; Africa [21%]), stunting (Asia [53%]; Africa [41%]), and overweight (Asia [48%]; Africa [27%]) [6,7]. In 2019, an estimated 47 million children under five were wasted, 38 million were overweight, and 144 million were stunted [8]. Sub-Saharan Africa alone accounts for nearly 58 million stunted children [9]. Unlike other regions, this number continues to rise, increasing from 49.7 million in 2000 to 57.5 million in 2019 [8]. These figures underscore persistent inequalities in access to adequate nutrition across low- and middle-income countries [10;9].

In Ghana, undernutrition remains a significant public health challenge. The Ghana Statistical Service [2023] reports that 17% of children under five are stunted, 6% wasted, and 12% underweight, with pronounced regional disparities [11; 12]. The Northern and Northeast regions record the highest stunting rates [29–30%], compared with 10–11% in the Eastern and Greater Accra regions [12]. In northern Ghana, up to 36% of children are stunted, 11% wasted, and 18% underweight [13]. Anaemia also remains prevalent, affecting nearly half of children aged 6–59 months [12; 14].

The consequences of undernutrition are profound, affecting growth, cognition, and future economic productivity and perpetuating intergenerational cycles of poverty [15;16]. Evidence highlights the importance of interventions during the first 1,000 days of life, when nutrition has the greatest influence on lifelong health outcomes [17–19].

In Ghana’s northern regions, research has identified maternal education, antenatal care, household wealth, sanitation, and family structure as significant determinants of child undernutrition [20]. Addressing child undernutrition requires multisectoral collaboration involving political actors, policymakers, civil society, academia, traditional and religious bodies, and development partners. Nutrition-specific and nutrition-sensitive interventions must prioritise high geographical coverage and integration across sectors [21]. Effective interventions such as community involvement, capacity building, and promoting nutrition-sensitive practices like the consumption of orange-fleshed sweet potatoes have successfully reduced malnutrition [22]. However, challenges such as sectoral agenda conflicts, technical skill gaps, and insufficient ownership hinder the delivery of multisectoral nutrition programs [23].

However, despite ongoing national strategies and interventions, there remains a limited understanding of how place-based factors, such as culture, environmental conditions, and community practices, influence child nutritional outcomes in rural Northeast Ghana. This gap highlights the need for region-specific evidence to guide effective, locally adapted interventions. Therefore, this study examines the prevalence and underlying determinants of child undernutrition in rural North-Eastern Ghana to inform context-appropriate policy and practice.

## Materials and methods

### Ethics Statement

Ethical approval for this study was obtained from the Kwame Nkrumah University of Science and Technology, Kumasi, Committee on Human Research, Publication and Ethics **(CHRPE/AP/149/23),** with permission from the East Mamprusi Municipal Health Directorate of the Ghana Health Service **(GHS/NER/EMM/10/2023).** The study was registered with the Pan African Clinical Trial Registry **(PACTR202412697952932).** Written informed consent was obtained from all participants, with explicit permission from parents/guardians for the participation of children. Participation was entirely voluntary, with no coercion, and participants could withdraw at any time without consequence. Confidentiality of all collected data was strictly maintained. Community entry procedures included meetings with traditional leaders and community representatives to ensure respect and cultural sensitivity throughout data collection and the entire study period.

### Study design and area

A community-based cross-sectional study was conducted in June/July 2023 to examine the prevalence and correlates of malnutrition and anaemia among children under five years in rural North-eastern Ghana for developing effective interventions. The data was collected in 10 selected rural communities across all the sub-districts within the East Mamprusi municipality, one of the six districts (***Figure 1***) of the Northeast Region of Ghana. According to the municipal health directorate’s 2020 annual report, the municipality comprises five (5) sub-districts, 241 operational communities, and 30 CHPS zones with services, 2 clinics, 4 health centres, and 1 hospital, which currently serves as the district hospital.

**Figure 1.**
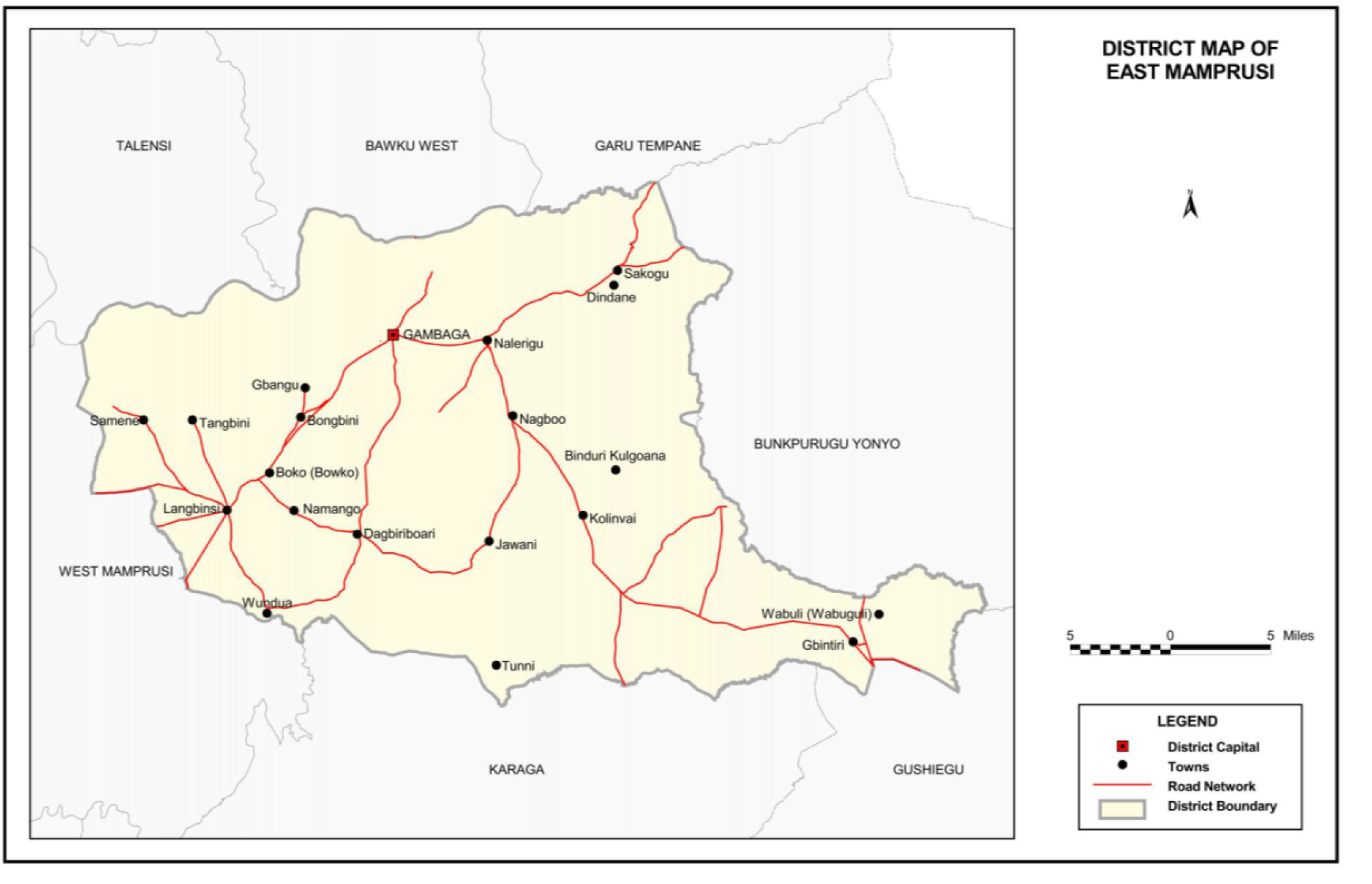
District map of East Mamprusi. *Source: Ghana Statistical Service, Population and Housing Census (2014)*.

### Sample size and recruitment

The initial sample size was estimated using G*Power 3.1.9.7 based on a two-tailed correlation (α = 0.05, power = 0.80, r = 0.30), which indicated that a minimum of 85 households would be required. Although this calculation was appropriate for detecting moderate associations, the study’s primary objectives to estimate the prevalence of household food insecurity and identify its correlates warranted a larger and more robust sample.

To better align with these aims, the sample was expanded to 248 households to ensure adequate power for logistic regression analyses examining associated factors of food insecurity and to account for potential missing data and subgroup analyses (e.g., by community or household characteristics). This larger sample size enhanced the precision of prevalence estimates and the stability of multivariable regression models.

A multistage cluster sampling strategy was utilised. Two rural communities were systematically selected from each of the five sub-districts of East Mamprusi Municipality (10 in total). Within these communities, the pen-spin method was used from a central point to identify households, and trained enumerators conducted household recruitment. Inclusion criteria required at least one child under five years old with a consenting primary caregiver; in cases with multiple eligible children, one was randomly selected per household. Children with acute illnesses were excluded and referred for care. This includes febrile illnesses (e.g., malaria, respiratory infections), diarrhoeal diseases, or any condition that causes significant discomfort, dehydration, or weakness at the time of data collection.

### Assessment of socio-demographic and socio-economic factors

Socio-demographic and socio-economic data were collected through one-on-one interviews with caregivers using a structured questionnaire. Child-related variables included date of birth and age in months, sex, and birth order. Maternal characteristics assessed were marital status, religion, ethnicity, educational attainment, occupation, parity, and household size. Socio-economic indicators such as estimated monthly expenditure on food for mother and child were also recorded.

### Measuring the Nutritional Status of Children

Trained anthropometrics took the weight and height/length for linear growth nutritional status assessment. Phlebotomists also collected serum blood, stool, and urine from study participants for examinations for potential parasites, sickling, G6PD, and full blood count.

### Linear growth assessment

Anthropometric measurements were conducted following standard protocols to ensure accuracy and reduce inter-observer variability [24]. Two trained data collectors independently measured each child, and discrepancies were resolved through repeated measurements. Weight was measured to the nearest 0.1 kg using a Seca 762 mechanical scale [Seca GmbH & Co. KG, Hamburg, Germany]. Children wore light clothing, and for infants or uncooperative children, maternal weight was taken together with the child and alone and used to calculate child weight by subtraction. Height or recumbent length was measured to the nearest 0.1 cm using a UNICEF height board. Recumbent length was recorded for children <24 months, while standing height was measured for those ≥24 months.

Child anthropometric indices, including weight-for-height [WHZ], weight-for-age [WAZ], and height-for-age [HAZ] z-scores, were computed using WHO Anthro Plus. Nutritional status categories were determined based on WHO cut-offs [Table 3.1] [World Health Organisation, 2017]. Additionally, the Cumulative Index of Anthropometric Failure [CIAF] was calculated to provide an overall measure of growth faltering, capturing children with any form of undernutrition [stunting, wasting, or underweight], thereby offering a comprehensive assessment of linear growth failure.

**Table 1:** WHO Nutritional status classification criteria.

| Nutritional Status | Indicator and cut-off value compared to the median |
| --- | --- |
| Overweight | Weight-for-length/height or BMI-for-age $> 2$ SD and $\leq 3$ SD of the median |
| Moderately underweight | Weight-for-age $\leq -2$ SD and $\geq -3$ SD of the median |
| Severely underweight | Weight-for-age $< -3$ SD of the median |
| Moderate acute malnutrition | Weight-for-length/height or BMI-for-age $\leq -2$ SD and $\geq -3$ SD of the median, or mid-upper arm circumference $\geq 115$ mm and $< 125$ mm |
| Severe acute malnutrition | Weight-for-length/height or BMI-for-age $< -3$ SD of the median or mid-upper arm circumference $< 115$ mm, or bilateral pitting oedema |
| Moderately stunted [moderate chronic malnutrition] | Length/height-for-age $\leq -2$ SD and $\geq -3$ SD of the median |
| Severely stunted [severe chronic malnutrition] | Length/height-for-age $< -3$ SD of the median |
| Moderately wasted | Weight-for-length/height $\leq -2$ SD and $\geq -3$ SD of the median |
| Severely wasted | Weight-for-length/height $< -3$ SD of the median |

[Source: World Health Organization, [24]

### Biochemical Assessment

Venous blood samples [6 mL] were collected by trained phlebotomists into silica-coated serum tubes [Becton Dickinson Vacutainer, Franklin Lakes, NJ, USA]. Haemoglobin [Hb] concentration and other haematological parameters were determined using full blood count analysis at the College of Nursing and Midwifery Laboratory, Nalerigu, Ghana. In addition, phlebotomists collected serum, stool, and urine samples from study participants. These were examined for intestinal parasites [stool microscopy], haematuria and urinary schistosomiasis [urine microscopy], sickle cell status [sickling test], and glucose-6-phosphate dehydrogenase [G6PD] deficiency [fluorescent spot test]. Anaemia was defined according to World Health Organisation thresholds [25]. Among children aged 6–59 months, anaemia severity was classified as: normal [≥11.0 g/dL], mild [10.0–10.9 g/dL], moderate [7.0–9.9 g/dL], and severe [<7.0 g/dL] [WHO, 2011; Domenica Cappellini & Motta, 2015]. Iron Deficiency Anaemia [IDA] was further screened using the Mentzer Index [MCV/RBC <13 suggestive of thalassaemia trait; >13 indicative of IDA]. The use of the Mentzer Index was therefore considered an appropriate proxy measure to estimate the burden of likely IDA within the available resources[25;26;27]

### Data Processing and Statistical Analysis

Data was collected using the KOBO Collect tool. The form was converted into a digital version of the questionnaire to integrate data collection and entry. Outliers and typographical errors were checked, and data were exported into SPSS version 24 for analysis. Nutritional status of participants was assessed using the WHO AnthroPlus software version 1.0.4, which converted anthropometric measurements into standardised z-scores: weight-for-height [WHZ], weight-for-age [WAZ], and height-for-age [HAZ]. These z-scores were imported into SPSS for further analysis. Nutritional status categories were determined using the WHO growth reference standards [26]

Descriptive statistics were used to summarise sociodemographic characteristics, dietary intake, and anthropometric indices. Associations between independent variables and child nutritional status indicators were examined using chi-square tests. Binary logistic regression was then applied to identify key associated factors of malnutrition [stunting, wasting, and underweight] and anaemia. Both crude odds ratios [COR] and adjusted odds ratios [AOR] with 95% confidence intervals [CI] were reported. Adjusted models accounted for the child’s age and sex, maternal education, maternal nutrition practices during pregnancy, diarrhoeal episodes, household wealth index, and household-level factors including sanitation and home health support. p-values <0.05 were considered statistically significant.

## Results

### Socio-Demographics of the Respondents

The study included 248 children aged 6–59 months, with a slightly higher proportion of males [56.0%] than females [44.0%]. Most mothers had no formal education [74.2%] and were engaged in farming [73.4%], while fathers were also predominantly farmers [87.1%]. The population was mainly Mamprusi [56.5%] and Komkomba [27.4%] by ethnicity, with Christianity [52.4%] and Islam [46.8%] as the dominant religions. Nearly all households were male-headed [98.8%]. Access to safe water was mostly through boreholes/pipe-borne sources [60.5%], and the majority of households used Kumasi Ventilated Improved Pit [KVIP] latrines [60.5%]. About 43% of children were served food in separate bowls [Table 2].

**Table 2:** Key Socio-Demographics of Study Population.

| **Variable** | **Frequency [n]** | **Percentage [%]** |
| --- | --- | --- |
| **Sex of child** |  |  |
| Male | 139 | 56.0 |
| Female | 109 | 44.0 |
| **Mother's education** |  |  |
| No formal education | 184 | 74.2 |
| Primary/JHS | 56 | 22.6 |
| SHS/Technical/Vocational | 8 | 3.2 |
| **Mother's occupation** |  |  |
| Farmer | 182 | 73.4 |
| Others [housewife/trader] | 66 | 26.6 |
| **Father's occupation** |  |  |
| Farmer | 216 | 87.1 |
| Others | 32 | 12.9 |
| **Ethnicity** |  |  |
| Mamprusi | 140 | 56.5 |
| Komkomba | 68 | 27.4 |
| Others | 40 | 16.1 |
| **Religion** |  |  |
| Christian | 130 | 52.4 |
| Muslim | 116 | 46.8 |
| Others | 2 | 0.8 |
| **Household head** |  |  |
| Male | 245 | 98.8 |
| Female | 3 | 1.2 |
| **Sanitation [toilet type]** |  |  |
| KVIP | 150 | 60.5 |
| Open defecation | 71 | 28.6 |
| Water closet | 27 | 10.9 |
| **Source of drinking water** |  |  |
| Borehole/pipe-borne | 150 | 60.5 |
| Open well | 71 | 28.6 |
| Stream/dam | 27 | 10.9 |
| **Child feeding practice** |  |  |
| Separate bowl | 106 | 42.7 |
| With mother/siblings | 142 | 57.3 |

### Child Nutrition Characteristics

The prevalence of linear growth malnutrition among children aged 6–59 months is presented in Table 3. Overall, stunting affected 34.3% of children, of which 19.0% were moderate and 15.3% were severe. Wasting was observed in 31.9% of children, with 18.5% moderate and 13.3% severe cases. Underweight prevalence was 27.8%, comprising 17.7% moderate and 10.1% severe. Thus, stunting was the most prevalent form of malnutrition, followed closely by wasting, while underweight was comparatively lower.

**Table 3:** Prevalence of Malnutrition among Children.

| **Stunting** | Frequency | Percent |
| --- | --- | --- |
| Normal | 163 | 65.7 |
| Moderate | 47 | 19.0 |
| Severe | 38 | 15.3 |
| **Wasting** |  |  |
| Normal | 169 | 68.1 |
| Moderate | 46 | 18.5 |
| Severe | 33 | 13.3 |
| **Underweight** |  |  |
| Normal | 179 | 72.2 |
| Moderate | 44 | 17.7 |
| Severe | 25 | 10.1 |

*[WHO Multicentre Growth Reference Study Group, 2006]; normal, moderate and severe Weight for Age Z-score [WAZ], Length for age Z-score [LAZ] and BMI for age [BMIAZ] are $-2 < Z\text{-score} < +2$ ; $-3 < Z\text{-score} < -2$ ; and $Z\text{-score} < -3$ , respectively.*

### Multiple index malnutrition

Table 4 illustrates that out of 248 children, 123 [49.6%] had no form of growth failure. 44 [17.7%], 54 [21.8%], and 27 [10.9%] of all children assessed suffer from single, double, or all three forms of anthropometric growth failure, manifesting as being underweight, stunting, and wasting in the same given children. In addition, 52 [21.0%], 50 [20.2%], and 94 [37.9%] of children had either stunting OR wasting, stunting OR underweight and wasting OR underweight, respectively. Also, double-index malnutrition co-occurrence in the same child showed that 56 [22.6%] were both wasted and stunted, 52 [21.0%] were both stunted and underweight, and 27 [10.9%] were both wasted and underweight

**Table 4:**
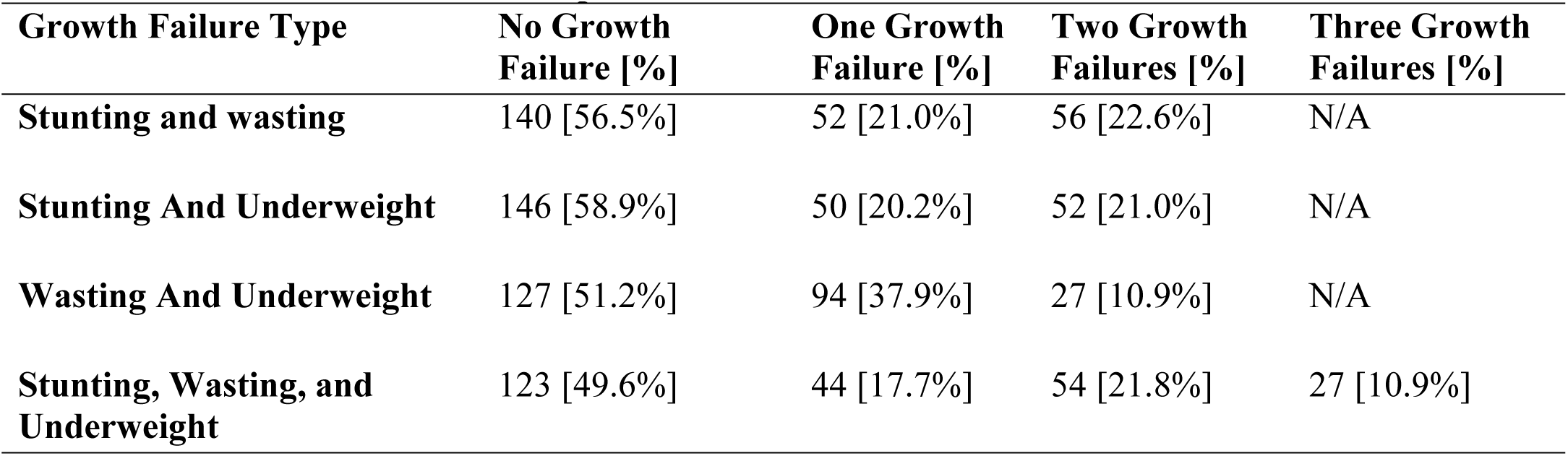
Cumulative Index of Anthropometric Growth Failure.

### Key Associated Factors of Stunting

Table 5 presents the factors associated with stunting in children through binary logistic regression. Children from households with higher estimated mother-child monthly food expenditure had reduced odds of stunting, with this association remaining statistically significant after adjustment for potential confounders [child’s age, sex, and maternal education] [AOR = 0.973, 95% CI: 0.967–0.979, p < 0.001]. Adequate knowledge of exclusive breastfeeding also showed a protective effect, remaining significant in the adjusted model [AOR = 0.38, 95% CI: 0.16–0.91, p = 0.030]. Similarly, crude analysis suggested that practising exclusive breastfeeding for six months reduced stunting risk, though this was not significant after adjustment [AOR = 0.53, 95% CI: 0.18–1.62, p = 0.266]. Knowledge about feeding sick children was unexpectedly associated with higher odds of stunting in the adjusted analysis [AOR = 3.96, 95% CI: 1.42–11.04, p = 0.009]. Adequate practice of Essential Nutrition Actions [ENAs] and meeting minimum dietary diversity scores were protective in crude models, but only dietary diversity remained significant after adjustment [AOR = 3.93, 95% CI: 1.51–10.19, p = 0.005]. Child platelet levels were also significant associated factors: normal platelet counts reduced the odds of stunting [AOR = 0.05, 95% CI: 0.00–0.82, p = 0.036], and low platelet levels were similarly protective [AOR = 0.02, 95% CI: 0.00–0.44, p = 0.013].

**Table 5.** Binary Logistic Regression for Factors Associated with Stunting.

| **Variables** | **Crude Odds Ratio [COR]**<br>**[95% CI]** | **p-value** | **Adjusted Odds Ratio [AOR]**<br>**[95% CI]** | **p-value** |
| --- | --- | --- | --- | --- |
| **Estimated Mother-Child monthly food expenditure** | 0.973 [0.967–0.979] | <0.001 | 0.973 [0.967–0.979] | <0.001 |
| **Knowledge of exclusive breastfeeding** |  |  |  |  |
| Adequate knowledge | 0.38 [0.14, 1.01] | 0.057 | 0.38 [0.16, 0.91] | 0.030 |
| Inadequate knowledge [*reference*] |  |  |  |  |
| **Practiced exclusive breastfeeding** |  |  |  |  |
| Practice 6 months of breastfeeding | 0.10 [0.05, 0.21] | <0.001 | 0.53 [0.18, 1.62] | 0.266 |
| Practice less than 6 months [*reference*] |  |  |  |  |
| **Knowledge on ACF** |  |  |  |  |
| Adequate knowledge | 0.19 [0.10, 0.34] | <0.001 | 2.05 [0.77, 5.47] | 0.153 |
| Inadequate knowledge [*reference*] |  |  |  |  |
| **Knowledge on feeding a sick child** |  |  |  |  |
| Adequate knowledge | 0.23 [0.14, 0.37] | <0.001 | 3.96 [1.42, 11.04] | 0.009 |
| Inadequate knowledge [*reference*] |  |  |  |  |
| **Knowledge of how to prevent IDA** |  |  |  |  |
| Adequate knowledge | 0.20 [0.12, 0.34] | <0.001 | 4.37 [1.75, 10.92] | 0.002 |
| Inadequate knowledge [*reference*] |  |  |  |  |
| **Knowledge on how to prevent IDD** |  |  |  |  |
| Adequate knowledge | 0.42 [0.24, 0.74] | <0.001 | 1.45 [0.52, 4.08] | 0.478 |
| Inadequate knowledge [*reference*] |  |  |  |  |
| **Overall practice of ENAs** |  |  |  |  |
| Adequate knowledge | 0.13 [0.07, 0.23] | <0.001 | 3.72 [0.90, 15.32] | 0.069 |
| Inadequate practice [*reference*] |  |  |  |  |
| **Minimum dietary diversity score category** |  |  |  |  |
| Met | 0.26 [0.15, 0.43] | <0.001 | 3.93 [1.51, 10.19] | 0.005 |
| Not met [*reference*] |  |  |  |  |
| **Child PLT FLAG [Interpretation]** |  |  |  |  |
| Normal | 0.25 [0.08, 0.74] | 0.013 | 0.05 [0.00, 0.82] | 0.036 |
| Low | 1.18 [0.62, 2.22] | 0.695 | 0.02 [0.00, 0.44] | 0.013 |
| High [*reference*] |  |  |  |  |

NOTE: ACF = Appropriate Complementary Feeding, IDA = Iron Deficiency Anaemia, IDD = Iodine Deficiency Disorders, ENAs = Essential Nutrition Actions, PLT FLAG = Platelet count interpretation. \*\* $p < .05$ indicates statistical significance.

### Key Factors Associated with Child Wasting

Table 6 shows that feeding practices were significantly associated with wasting. Children who were served meals in a separate bowl had a markedly reduced likelihood of being wasted compared to those who ate with their siblings [AOR = 0.057; 95% CI: 0.014–0.237; *p* < 0.001]. Children who were not introduced to appropriate complementary feeding [ACF] were over 11 times more likely to be wasted than those who received ACF [AOR = 11.231; 95% CI: 2.347–53.744; *p* = 0.002].

**Table 6:** Binary Logistic Regression for Factors Associated with Child Wasting.

| Variables | Crude odds ratio [COR] [95% CI] | P-value [COR] | Adjusted odds ratio [AOR] [95% CI] | P-value [AOR] |
| --- | --- | --- | --- | --- |
| **How a child is served** |  |  |  |  |
| Separate bowl | 0.057 [0.014, 0.237] | <0.001 | 0.057 [0.014, 0.237] | <0.001 |
| With mother | 0.311 [0.079, 1.224] | 0.095 | 0.311 [0.079, 1.224] | 0.095 |
| With siblings [*reference*] |  |  |  |  |
| **Practiced exclusive breastfeeding** |  |  |  |  |
| No | 0.406 [0.094, 1.756] | 0.228 | 0.406 [0.094, 1.756] | 0.228 |
| Yes [*reference*] |  |  |  |  |
| **Practiced ACF** |  |  |  |  |
| No | 11.231 [2.347, 53.744] | 0.002 | 11.231 [2.347, 53.744] | 0.002 |
| Yes [*reference*] |  |  |  |  |
| **Practiced nutrition during pregnancy** |  |  |  |  |
| No | 0.198 [0.059, 0.666] | 0.009 | 0.198 [0.059, 0.666] | 0.009 |
| Yes [*reference*] |  |  |  |  |
| **Practiced prevention of vitamin A deficiency** |  |  |  |  |
| No | 13.241 [0.928, 189.004] | 0.057 | 13.241 [0.928, 189.004] | 0.057 |
| Yes [*reference*] |  |  |  |  |
| **Practiced feeding a sick child** |  |  |  |  |
| No | 1.518 [0.348, 6.624] | 0.578 | 1.518 [0.348, 6.624] | 0.578 |
| Yes [*reference*] |  |  |  |  |
| **Practiced prevention of IDA** |  |  |  |  |
| No | 5.654 [1.708, 18.715] | 0.005 | 5.654 [1.708, 18.715] | 0.005 |
| Yes [*reference*] |  |  |  |  |
| **Practiced prevention of IDD** |  |  |  |  |
| No | 8.371 [1.934, 36.236] | 0.004 | 8.371 [1.934, 36.236] | 0.004 |
| Yes [*reference*] |  |  |  |  |
| **Overall ENAs practiced category** |  |  |  |  |
| Adequate practice | 0.385 [0.038, 3.907] | 0.419 | 0.385 [0.038, 3.907] | 0.419 |
| inadequate practice [*reference*] |  |  |  |  |
| **Minimum dietary diversity score** |  |  |  |  |
| Not met | 2.211 [0.636, 7.693] | 0.212 | 2.211 [0.636, 7.693] | 0.212 |
| Met [*reference*] |  |  |  |  |

*Note. p < .05 indicates statistical significance.*

Maternal nutrition practices also showed significant associations. Children whose mothers did not practice good nutrition during pregnancy had significantly higher odds of wasting [AOR = 0.198; 95% CI: 0.059–0.666; *p* = 0.009]. In addition, the lack of prevention of iron deficiency anaemia [IDA] increased the odds of wasting by more than five-fold [AOR = 5.654; 95% CI: 1.708–18.715; *p* = 0.005]. Similarly, children whose caregivers did not practice prevention of iodine deficiency disorders [IDD] had over eight times higher odds of wasting compared to those who did [AOR = 8.371; 95% CI: 1.934–36.236; *p* = 0.004].

Other variables, including exclusive breastfeeding, prevention of vitamin A deficiency, feeding during illness, overall Essential Nutrition Actions [ENAs] practice category, and minimum dietary diversity score, were not significantly associated with child wasting [*p* > 0.05].

### Key Factors Associated with Child Underweight

The binary logistic regression analysis results in Table 7 indicate that lack of home support from healthcare providers was strongly associated with child underweight, as children whose households did not receive home support were over 21 times more likely to be underweight compared with those who received support [AOR = 21.443; 95% CI: 3.386–135.775; *p* = 0.001]. Feeding practices were also influential. Children who were not exclusively breastfed had significantly higher odds of being underweight [AOR = 9.530; 95% CI: 1.712–53.058; *p* = 0.010]. Similarly, children whose caregivers did not practice appropriate feeding during illness were about ten times more likely to be underweight [AOR = 10.703; 95% CI: 1.804–63.506; *p* = 0.009].

**Table 7:**
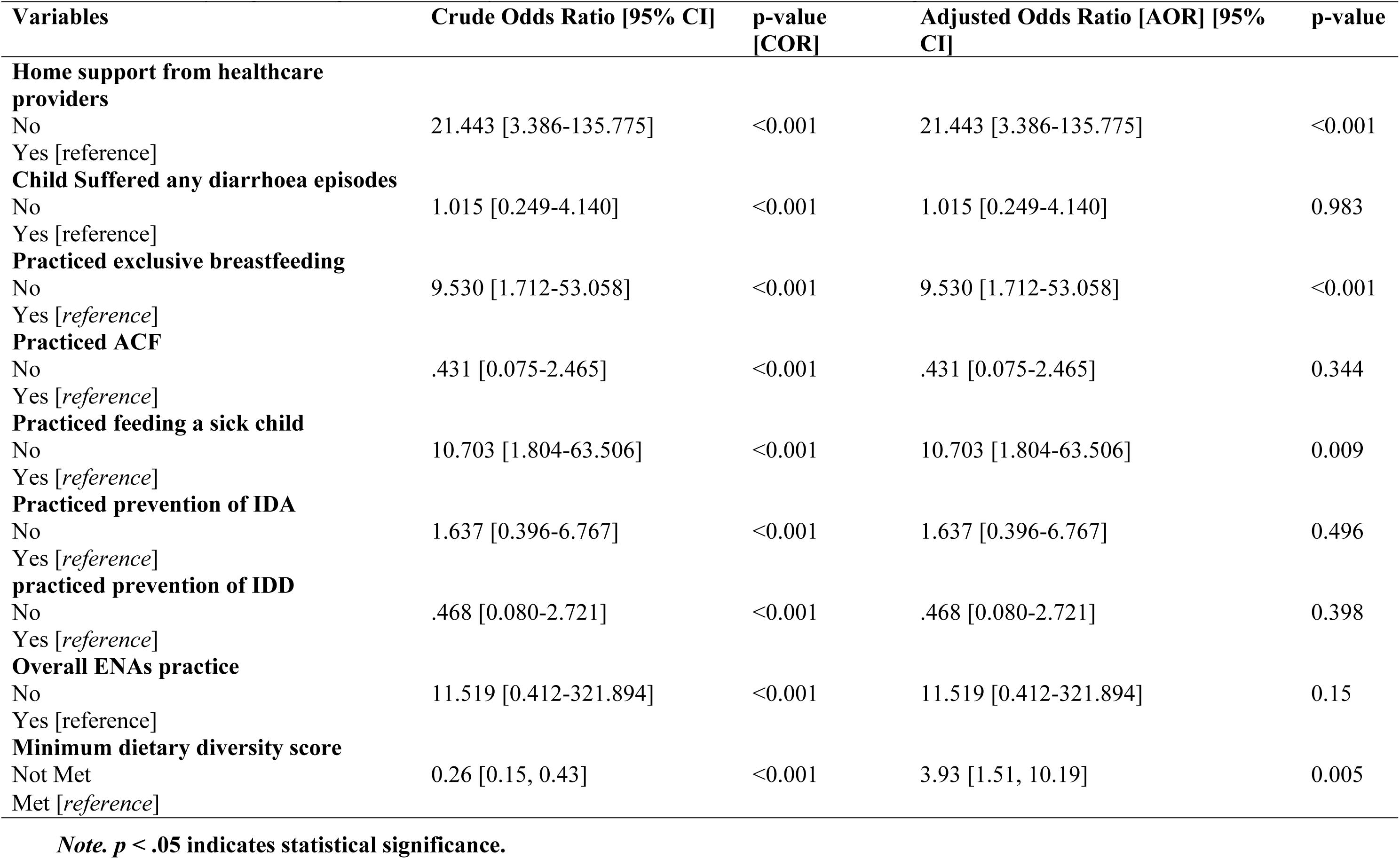
Binary Logistic Regression for Key Factors Associated with Child Underweight.

| **Variables** | **Crude Odds Ratio [95% CI]** | **p-value [COR]** | **Adjusted Odds Ratio [AOR] [95% CI]** | **p-value** |
| --- | --- | --- | --- | --- |
| **Home support from healthcare providers** |  |  |  |  |
| No | 21.443 [3.386-135.775] | <0.001 | 21.443 [3.386-135.775] | <0.001 |
| Yes [reference] |  |  |  |  |
| **Child Suffered any diarrhoea episodes** |  |  |  |  |
| No | 1.015 [0.249-4.140] | <0.001 | 1.015 [0.249-4.140] | 0.983 |
| Yes [reference] |  |  |  |  |
| **Practiced exclusive breastfeeding** |  |  |  |  |
| No | 9.530 [1.712-53.058] | <0.001 | 9.530 [1.712-53.058] | <0.001 |
| Yes [reference] |  |  |  |  |
| **Practiced ACF** |  |  |  |  |
| No | .431 [0.075-2.465] | <0.001 | .431 [0.075-2.465] | 0.344 |
| Yes [reference] |  |  |  |  |
| **Practiced feeding a sick child** |  |  |  |  |
| No | 10.703 [1.804-63.506] | <0.001 | 10.703 [1.804-63.506] | 0.009 |
| Yes [reference] |  |  |  |  |
| **Practiced prevention of IDA** |  |  |  |  |
| No | 1.637 [0.396-6.767] | <0.001 | 1.637 [0.396-6.767] | 0.496 |
| Yes [reference] |  |  |  |  |
| **practiced prevention of IDD** |  |  |  |  |
| No | .468 [0.080-2.721] | <0.001 | .468 [0.080-2.721] | 0.398 |
| Yes [reference] |  |  |  |  |
| **Overall ENAs practice** |  |  |  |  |
| No | 11.519 [0.412-321.894] | <0.001 | 11.519 [0.412-321.894] | 0.15 |
| Yes [reference] |  |  |  |  |
| **Minimum dietary diversity score** |  |  |  |  |
| Not Met | 0.26 [0.15, 0.43] | <0.001 | 3.93 [1.51, 10.19] | 0.005 |
| Met [reference] |  |  |  |  |

*Note.* $p < .05$ indicates statistical significance.

Dietary diversity showed a significant effect as children who did not meet the minimum dietary diversity score were almost four times more likely to be underweight compared to those who met the requirement [AOR = 3.93; 95% CI: 1.51–10.19; *p* = 0.005].

Other factors, including serving style [separate bowl vs. with others], maternal knowledge of nutrition during pregnancy, appropriate complementary feeding [ACF], prevention of iron deficiency anaemia [IDA], prevention of iodine deficiency disorders [IDD], and overall Essential Nutrition Actions [ENA] practice were not significantly associated with child underweight after adjustment [*p* > 0.05].

### Correlations of Nutritional Status

Pearson correlation analyses [Table 8, Appendix I] revealed significant associations between household food expenditure, maternal characteristics, and child nutritional indicators. A strong positive correlation was observed between estimated monthly food expenditure and child height-for-age Z-score [r = 0.714, p < 0.001]. Food expenditure was also moderately and positively correlated with weight-for-height Z-score [r = 0.491, p < 0.001] and weight-for-age Z-score [r = 0.433, p < 0.001]. In addition, food expenditure showed a modest but significant positive association with child haemoglobin levels [r = 0.242, p < 0.001]. Maternal age demonstrated weak positive associations with child haemoglobin level [r = 0.142, p = 0.025]. No significant correlations were found between maternal age and weight-for-height Z-score [r = 0.013, p = 0.835] or weight-for-age Z-score [r = 0.038, p = 0.546].

Child haemoglobin status was modestly correlated with both weight-for-height Z-score [r = 0.165, p = 0.009] and weight-for-age Z-score [r = 0.195, p = 0.002].

In contrast, the minimum dietary diversity score was not significantly associated with any child anthropometric indicator, including height-for-age [r = 0.034, p = 0.591], weight-for-height [r = 0.029, p = 0.650], or weight-for-age [r = 0.056, p = 0.377]. Similarly, no significant relationships were observed between the Household Food Insecurity Access Score and child haemoglobin [r = 0.047, p = 0.458].

### Haematological Profile and Anaemia Prevalence in Children

Results presented in Table 9 assessed glucose-6-phosphate dehydrogenase [G6PD] deficiency, sickling status, parasitic infections, and anaemia prevalence. Most children [58.5%, n=145] had normal G6PD status, while 24.6% [n=61] showed G6PD deficiency and 16.9% [n=42] had partial deficiency. Sickling disease prevalence was low, with only 4.8% [n=12] testing positive. No parasitic infections were detected in the study population.

**Table 9.** Frequency and Percentage Distribution of Haematological and Anaemia Status.

| Variable | Category | Frequency [n] | Percent [%] |
| --- | --- | --- | --- |
| **G6PD Status** | No Defect | 145 | 58.5 |
|  | Partial | 42 | 16.9 |
|  | Defective | 61 | 24.6 |
| **Sickling Status** | Negative | 236 | 95.2 |
|  | Positive | 12 | 4.8 |
| **Parasite Flag** | Not Seen | 248 | 100.0 |
| **Anaemia Status** | Not Anaemic | 100 | 40.3 |
|  | Mild | 56 | 22.6 |
|  | Moderate | 84 | 33.9 |
|  | Severe | 8 | 3.2 |
| **Anaemia Type** | Non-anaemic | 100 | 40.3 |
|  | IDA | 137 | 55.2 |
|  | Other Forms | 11 | 4.4 |

*Anaemia classification based on WHO guidelines: Normal $\geq 11.0$ g/dl, Mild 10.0–10.9 g/dl, Moderate 7.0–9.9 g/dl, Severe $< 7.0$ g/dl [ WHO, 2011; Domenica Cappellini & Motta, 2015]. **Mentzer Index** = Mean Corpuscular Volume [MCV] /the Red Blood Cell [RBC] count $< 13$ : Suggests Thalassemia. **Mentzer Index** $> 13$ : Suggests Iron Deficiency Anaemia [IDA]. [27]*

Anaemia affected 59.7% [n=148] of children, with moderate anaemia being the most prevalent form [33.9%, n=84], followed by mild anaemia [22.6%, n=56], while severe anaemia was observed in only 3.2% [n=8] of participants. Among anaemic children, iron deficiency anaemia [IDA] was predominant, affecting 55.2% [n=137] of the total sample, as determined by Mentzer Index analysis. Other forms of anaemia were uncommon, accounting for only 4.4% [n=11] of cases.

### Factors Associated with Childhood Anaemia

Chi-square analysis identified several factors significantly associated with anaemia status among children [Table 10, Appendix I]. Anaemia prevalence was higher among children whose mothers were unsatisfied with maternal health services [77.4% vs. 57.1%, *p* = 0.031] and who reported poor service quality [88.2% vs. 54.1%, *p* = 0.001]. Children whose mothers had not undergone haemoglobin checks at antenatal registration were more likely to be anaemic [78.8% vs. 56.7%, *p* = 0.016].

Lack of home support from healthcare workers after birth was strongly associated with anaemia [75.8% vs. 49.0%, *p* < 0.001], as was the absence of a child health record book [87.5% vs. 57.1%, *p* = 0.049]. Children living in nuclear households had a higher prevalence of anaemia compared to those living in extended households [66.1% vs. 53.8%, *p* = 0.049].

Anaemia was significantly more prevalent among children with diarrhoea [80.0% vs. 55.2%, *p* = 0.002], those not exclusively breastfed [79.2% vs. 50.9%, *p* < 0.001], not given adequate complementary feeding [68.3% vs. 53.5%, *p* = 0.019], or not fed during illness [68.3% vs. 51.6%, *p* = 0.007]. Children whose mothers did not practice iron deficiency anaemia prevention had markedly higher anaemia rates [85.5% vs. 39.1%, *p* < 0.001].

Growth indicators also showed strong associations: anaemia was more common in stunted [75.3% vs. 51.5%, *p* < 0.001] and wasted [77.2% vs. 51.5%, *p* < 0.001] children. Finally, maternal anaemia was associated with a higher prevalence of childhood anaemia [67.8% vs. 46.9%, *p* = 0.001].

### Factors associated with Child Anaemia

Binary logistic regression analysis was performed to identify key factors associated with childhood anaemia (Table 11). Having maternal haemoglobin checked at registration was strongly protective in both crude [COR = 0.168, 95% CI: 0.046–0.617, *p* = 0.006] and adjusted models [AOR = 0.132, 95% CI: 0.034–0.504, *p* = 0.003]. Maternal satisfaction with healthcare services and perceived service quality were significant in crude analyses but lost significance after adjustment. Similarly, possession of an original child health record showed reduced odds of anaemia in crude analysis but not after adjustment.

**Table 11:** Binary Logistic Regression Analyses of Factors Associated with Child Anaemia.

| Variable | Crude Odds Ratio [COR] | 95% CI [COR] | p-value [COR] | Adjusted Odds Ratio [AOR] | 95% CI | p-value |
| --- | --- | --- | --- | --- | --- | --- |
| **Satisfied with the maternal health services** |  |  |  |  |  |  |
| Yes | 0.265 | 0.092 - 0.757 | 0.012 | 0.341 | 0.088 - 1.326 | 0.12 |
| No [*reference*] |  |  |  |  |  |  |
| **Self-reported rating of quality of maternal health services** |  |  |  |  |  |  |
| Poor | 2.7 | 1.158 - 6.317 | 0.021 | 3.753 | 0.701 - 20.082 | 0.122 |
| Good | 2.2 | 0.755 - 6.306 | 0.145 | 3.88 | 0.560 - 26.863 | 0.17 |
| Excellent [*Reference*] |  |  |  |  |  |  |
| **Hb checked at registration** |  |  |  |  |  |  |
| Yes | 0.168 | 0.046 - 0.617 | 0.006 | 0.132 | 0.034 - 0.504 | 0.003 |
| No [*reference*] |  |  |  |  |  |  |
| **Type of child health record book** |  |  |  |  |  |  |
| Original | 0.263 | 0.079 - 0.871 | 0.028 | 0.369 | 0.026 - 5.309 | 0.463 |
| Improvised | 1.269 | 0.366 - 4.368 | 0.702 | 1.191 | 0.240 - 5.905 | 0.83 |
| No book [*Reference*] |  |  |  |  |  |  |
| **Practiced exclusive breastfeeding** |  |  |  |  |  |  |
| Yes | 0.309 | 0.104 - 0.927 | 0.036 | 0.311 | 0.088 - 1.098 | 0.069 |
| No [*reference*] |  |  |  |  |  |  |
| **Practiced on ACF** |  |  |  |  |  |  |
| Yes | 4.347 | 1.241 - 15.351 | 0.022 | 6.409 | 1.311 - 31.323 | 0.022 |
| No [*reference*] |  |  |  |  |  |  |
| **Practiced prevention of IDA** |  |  |  |  |  |  |
| Yes | 0.047 | 0.014 - 0.161 | 0.002 | 0.066 | 0.023 - 0.188 | <0.001 |
| No [*reference*] |  |  |  |  |  |  |
| **Mother HB binary** |  |  |  |  |  |  |
| Anaemia | 0.387 | 0.232 - 0.638 | <0.001 | 0.813 | 0.325 - 2.037 | 0.659 |
| Normal [*reference*] |  |  |  |  |  |  |

*Note.* $p < .05$ indicates statistical significance.

Exclusive breastfeeding reduced the odds of anaemia in crude analysis [COR = 0.309, 95% CI: 0.104–0.927, *p* = 0.036], with a similar but non-significant trend in the adjusted model [AOR = 0.311, 95% CI: 0.088–1.098, *p* = 0.069]. In contrast, children whose mothers reported practising appropriate complementary feeding [ACF] had significantly higher odds of anaemia [AOR = 6.409, 95% CI: 1.311–31.323, *p* = 0.022]. Preventive practices against iron deficiency anaemia [IDA] were strongly protective, reducing the odds by over 90% [AOR = 0.066, 95% CI: 0.023–0.188, *p* < 0.001].

Maternal anaemia status was significant in crude analysis [COR = 0.387, 95% CI: 0.232–0.638, *p* < 0.001], but this association disappeared in the adjusted model [AOR = 0.813, 95% CI: 0.325–2.037, *p* = 0.659].

## Discussion

This study provides an assessment of multiple forms of malnutrition and anaemia in an understudied rural population, using standardised WHO criteria and validated assessment tools. The findings revealed alarming rates of malnutrition among children aged 6-59 months in rural north-eastern Ghana, with 34.3% stunted, 31.9% wasted, and 27.8% underweight. The cumulative anthropometric growth failure index revealed that 50.4% of children experienced at least one form of malnutrition, indicating pervasive nutritional challenges within this population.

These figures are far above the national averages reported by GSS, GHS and ICF [12];18% stunting, 6% wasting, and 12% underweight, highlighting regional disparities. National-level studies corroborate these findings, showing that 35.6% of Ghanaian children under five experience some form of malnutrition, with 27.5% stunted, 13.8% underweight, and 8.9% wasted [28], while earlier data from 2008 indicated similarly high rates [29]. Using the extended composite index of anthropometric failure, estimates suggest that between one-third and half of all Ghanaian children aged 6–59 months suffer at least one form of malnutrition [30]. Rural communities are particularly vulnerable, as seen in Gbeo, north-eastern Ghana, where 34.4% of children were stunted, 18.8% underweight, and 37.9% wasted [31]. Such evidence reinforces that, although national progress has been made in reducing undernutrition, malnutrition remains disproportionately concentrated in rural and deprived areas. Targeted, context-specific interventions are therefore urgently needed to address these inequities and to support Ghana’s progress toward achieving both national and global nutrition targets.

The most significant associated factors across all forms of malnutrition was household food expenditure, with higher estimated monthly food expenditure strongly associated with reduced odds of stunting [AOR = 0.973, 95% CI: 0.967-0.979, p < 0.001] and showing strong positive correlations with all anthropometric indicators [height-for-age Z-score: r = 0.714, p < 0.001; weight-for-height Z-score: r = 0.491, p < 0.001; weight-for-age Z-score: r = 0.433, p < 0.001]. This finding underscores the crucial role of economic stability in nutritional outcomes, as supported by research across multiple countries, which shows that household food expenditure patterns significantly predict child growth outcomes [34; 33;32].

Feeding practices emerged as critical modifiable factors. For stunting, adequate knowledge of exclusive breastfeeding demonstrated protective effects [AOR = 0.38, 95% CI: 0.16-0.91, p = 0.030], while meeting minimum dietary diversity requirements paradoxically showed increased odds in the adjusted model [AOR = 3.93, 95% CI: 1.51-10.19, p = 0.005]. For wasting, children served meals in separate bowls had a markedly reduced likelihood of being wasted compared to those eating with siblings [AOR = 0.057, 95% CI: 0.014-0.237, p < 0.001], while lack of appropriate complementary feeding increased wasting odds over eleven-fold [AOR = 11.231, 95% CI: 2.347-53.744, p = 0.002]. Similarly, for underweight, exclusive breastfeeding failure significantly increased risk [AOR = 9.530, 95% CI: 1.712-53.058, p = 0.010]. Healthcare service utilisation proved crucial, particularly in preventing underweight, where a lack of home support from healthcare providers was associated with over 21 times higher odds of underweight [AOR = 21.443, 95% CI: 3.386-135.775, p = 0.001]. These findings align with extensive literature demonstrating that feeding practices are critical determinants of child nutritional status, with exclusive breastfeeding consistently emerging as a protective factor against stunting, wasting, and underweight across multiple studies [38; 37 36; 35;34].

Several associations observed in this study appeared paradoxical. For example, caregivers with adequate knowledge on feeding a sick child had higher odds of stunting, and meeting minimum dietary diversity was associated with increased odds of both stunting and underweight. Similarly, reported appropriate complementary feeding practices appeared linked with greater odds of anaemia. These counter-intuitive results likely reflect a combination of residual confounding, reverse causality, and contextual limitations. It is plausible that caregivers of already malnourished children may have acquired greater nutritional knowledge or improved feeding practices after their children were diagnosed with malnutrition, resulting in a misleading positive association. Measurement errors may also have contributed, as the knowledge and practice variables relied on self-reported responses, which may not accurately capture the depth, consistency, or accuracy of actual practices. Moreover, in rural contexts with constrained food environments, caregivers may meet dietary diversity using available but nutrient-poor or contaminated foods, thereby fulfilling criteria without achieving adequate nutrient intake. These highlight that knowledge and reported practices alone may not suffice to improve nutritional outcomes unless accompanied by structural improvements in food quality, accessibility, and household resources.

The heightened risk of malnutrition associated with shared feeding practices can be attributed to several factors. Sharing food from the same bowl limits individual control over portion sizes and nutrient intake, potentially resulting in some children receiving inadequate food or nutrients due to competition among siblings [39]. This practice also increases the risk of infection transmission, such as diarrhoea, which impairs nutrient absorption and exacerbates malnutrition [40]. Furthermore, shared feeding reflects less tailored feeding practices that do not accommodate the differing dietary needs or appetites of individual children, critical for proper growth and nutrition, especially in early childhood [41]. Additionally, shared feeding may reduce maternal attention to individual children’s intake and hygiene, particularly in large households, further contributing to nutritional inequities among siblings. Serving meals in separate bowls promotes fair food allocation, reduces contamination, and encourages responsive feeding, all crucial for growth and disease prevention [42].

Healthcare service utilisation, including postnatal home visits and healthcare provider support, in preventing underweight aligns with findings that adequate antenatal care attendance and maternal health-seeking behaviours are strongly associated with improved nutritional outcomes and reduced malnutrition prevalence [35;43;13].

Micronutrient deficiency prevention also emerged as crucial across all malnutrition forms. Lack of iron deficiency anaemia [IDA] prevention increased wasting odds five-fold [AOR = 5.654, 95% CI: 1.708-18.715, p = 0.005], while inadequate iodine deficiency disorder [IDD] prevention increased wasting odds eight-fold [AOR = 8.371, 95% CI: 1.934-36.236, p = 0.004]. These findings align with the Ghana Micronutrient Survey, which highlighted that micronutrient malnutrition, including iron and iodine deficiencies, continues to significantly affect children under five and contribute to wasting and other forms of undernutrition [40]. The survey also emphasised the importance of integrated, multi-nutrient approaches, including supplementation, fortification, and dietary diversification, to effectively prevent micronutrient deficiencies and reduce wasting among children in Ghana [40]. These results underscore the critical importance of integrated strategies addressing multiple micronutrient deficiencies simultaneously to improve child nutritional outcomes and health.

The prevalence of anaemia among children in this study was alarmingly high at 59.7%, with iron deficiency anaemia [IDA] constituting the majority at 55.2%. This is consistent with previous research indicating that anaemia affects approximately 62% of children under five in Ghana, and underscores the persistent challenge of micronutrient malnutrition, particularly iron deficiency [45]. A national survey found that 35.6% of children had anaemia, 21.5% had iron deficiency, and 12.2% had iron deficiency anaemia(42). It is important to acknowledge, however, that IDA classification in this study was based on the Mentzer Index, which, while practical in field settings, is primarily a screening tool rather than a diagnostic standard. Its reliance on red cell indices may lead to misclassification, especially in populations where thalassaemia and other haemoglobinopathies are present. Ideally, serum ferritin or transferrin receptor assays would provide stronger confirmation, but such analyses were not feasible due to resource constraints. Nevertheless, the high prevalence observed aligns with national evidence and highlights the urgent need for strengthened micronutrient interventions.

Historical studies show variability in prevalence estimates; for example, (43) reported a higher prevalence of 78.4%, while [46] documented a lower national prevalence of 35.6% for anaemia and 21.5% specifically for iron deficiency among children aged 6–59 months. The differences observed between these studies and the current finding of 55.2% IDA prevalence may be explained by variations in sample populations, geographical coverage, diagnostic criteria, temporal changes, and interventions implemented over time. Regional disparities, seasonal timing of surveys, and differences in laboratory methods for diagnosing iron deficiency can also contribute to such discrepancies. Nonetheless, the persistently high burden of anaemia, especially iron deficiency anaemia, underscores the urgent need for targeted nutritional and public health interventions in Ghana to improve child health outcomes. The interconnected nature of malnutrition and anaemia was evident, with anaemia significantly more common among stunted [75.3% vs. 51.5%, p < 0.001] and wasted children [77.2% vs. 51.5%, p < 0.001]. Studies consistently show higher anaemia prevalence among malnourished children, with stunted children having significantly elevated odds of anaemia [47; 48]. Key factors associated with childhood anaemia in this study included maternal healthcare practices, with haemoglobin checks at registration showing strong protective effects [AOR = 0.132, 95% CI: 0.034-0.504, p = 0.003]. This corresponds with Sowe et al. [49], study found that maternal healthcare practices emerge as crucial protective factors, with haemoglobin monitoring and iron deficiency anaemia prevention reducing childhood anaemia odds by over 90%. Preventive practices against IDA were highly protective, reducing anaemia odds by over 90% [AOR = 0.066, 95% CI: 0.023-0.188, p < 0.001]. The strong association between maternal and child anaemia status [67.8% vs. 46.9%, p = 0.001] emphasises the interconnectedness of maternal and child health, consistent with studies showing anaemic mothers are more likely to have anaemic children [49;50].

Micronutrient deficiency prevention emerged as crucial across all forms of malnutrition, as the results highlighted significant associations between the lack of iron deficiency anaemia [IDA] and iodine deficiency disorder [IDD] prevention, with increased malnutrition risk. This finding was confirmed by a report by [51], which indicates that IDA, IDD, vitamin A, zinc, and folate deficiencies are the most contributing to poor growth.

Micronutrient deficiencies, including iron, iodine, vitamin A, zinc, and folate, impair immune function, growth, and neurodevelopment, increasing vulnerability to infections and worsening malnutrition [52].

Iron deficiency impairs haemoglobin synthesis, leading to reduced oxygen transport to tissues, which compromises energy metabolism vital for physical growth and development [52;53] Chronic iron deficiency in infancy is associated with poorer cognitive function, motor development, and long-term neurodevelopmental deficits [53]. R. Bailey et al., [52] report that iodine deficiency results in mental retardation and reduced cognitive function, while Elizabeth L. Prado et al., [53] demonstrates that nutrient deficiencies can permanently impair neurodevelopmental processes. A. Imdad et al., [54] notes that micronutrient deficiencies are associated with 35% of child deaths under 5 years, with vitamin A, iron, zinc, and iodine being most prevalent. P. Bhaskaram et al.,[55] further emphasizes that these micronutrients have immunomodulating functions that influence disease susceptibility, creating a vicious cycle of malnutrition and infection.

This biological interdependence emphasises that interventions must be integrated, addressing multiple nutrient gaps simultaneously rather than focusing on single deficiencies.

Thus, these micronutrients are indispensable for multiple physiological and developmental processes. Their deficiencies disrupt normal growth trajectories, increasing the risk of wasting and other forms. This biological rationale emphasizes the importance of integrated nutrition interventions targeting multiple micronutrient deficiencies simultaneously to effectively improve child health outcomes and reduce malnutrition burdens.

The study population reflected significant socioeconomic challenges, with 74.2% of mothers lacking formal education and 73.4% engaged in farming. Access to essential services remained limited, with 28.6% of households practising open defecation and similar proportions relying on open wells for drinking water. These infrastructural deficits align with national challenges, where sanitation coverage lags at only 18.5% despite improved water access reaching 92.7% [54]

Community-level variations in malnutrition prevalence were evident, emphasising localised disparities in resource distribution and healthcare delivery. Traditional feeding practices, where 57.3% of children shared bowls with mothers or siblings, contextualise nutritional risks and highlight the need for culturally appropriate interventions. Recent studies in Ghana reveal similar prevalence variations of childhood malnutrition, ranging from 31.2% to 57.3% using composite anthropometric indices [30].

Overall, this study reveals that while maternal knowledge and feeding practices are crucial, they are insufficient in isolation. Nutritional knowledge must be supported by access to quality foods, healthcare, and improved sanitation to translate into measurable child health gains. Future longitudinal and mixed-method studies should explore how caregiver knowledge interacts with food availability, maternal autonomy, and household resources to shape nutritional outcomes.

In rural northeastern Ghana, the rates of child malnutrition and anaemia remain unacceptably high. Strengthening maternal education, improving food security, promoting hygienic feeding practices, and expanding healthcare access are essential for sustainable progress. Codesigned, community-based, and culturally grounded approaches are needed to ensure interventions are relevant, acceptable, and effective in reducing child malnutrition.

## Conclusion

This study highlights a persistently high burden of malnutrition and anaemia among children aged 6–59 months in rural north-eastern Ghana, revealing deep-seated nutritional and socioeconomic disparities. Stunting, wasting, and underweight remain widespread, compounded by a high prevalence of iron deficiency anaemia. The findings demonstrate that child nutrition is shaped by interconnected factors including household food expenditure, maternal education, feeding practices, healthcare utilisation, and micronutrient deficiency prevention.

Based on the strongest findings, several targeted interventions are recommended. First, promoting individual bowl feeding for young children should be prioritised to ensure equitable food distribution, reduce contamination, and support responsive feeding practices. Second, strengthening home-based healthcare and postnatal follow-up including home visits and counselling by trained community health workers—can significantly reduce underweight and improve overall nutritional outcomes. Third, routine maternal haemoglobin checks during antenatal and postnatal care should be reinforced to prevent both maternal and child anaemia, given the strong association between maternal and child haemoglobin status.

Addressing food insecurity through livelihood support, social protection schemes, and improved dietary diversity using locally available, nutrient-dense foods is also essential. These measures should be coupled with community nutrition education that moves beyond knowledge transfer to empower mothers and caregivers to translate knowledge into effective feeding and health-seeking practices.

Finally, codesigned, community-specific, and culturally appropriate interventions that engage local leaders, health providers, and caregivers are critical for sustainability. Such participatory approaches ensure that programmes reflect local realities, promote ownership, and enhance the likelihood of long-term success in reducing child malnutrition and anaemia.

## Study Limitations

This study has several limitations that should be acknowledged. The cross-sectional design limits causal inference, as it captures associations at a single point in time rather than establishing temporal relationships. Additionally, reliance on self-reported information from caregivers introduces potential recall bias, particularly regarding dietary practices and the use of health services. The study was also conducted exclusively in rural communities, which may limit the generalizability of the findings to urban settings or other regions in Ghana.

Furthermore, while the Mentzer Index was used to screen for Iron Deficiency Anaemia [IDA], it is primarily a practical screening tool rather than a definitive diagnostic method. Its reliance on red blood cell indices may lead to misclassification, especially in populations where thalassaemia and other haemoglobinopathies are prevalent. Ideally, confirmation through biochemical markers such as serum ferritin, transferrin saturation, or soluble transferrin receptor would provide a more accurate diagnosis. However, due to resource and logistical constraints in the study setting, such confirmatory analyses could not be conducted. Future longitudinal studies incorporating diverse populations, additional biomarkers, and broader geographical coverage are recommended to enhance causal interpretation and improve diagnostic accuracy.

## Declarations

### Ethics approval and consent to participate

The study obtained ethical clearance from the Kwame Nkrumah University of Science and Technology Committee on Human Research, Publications and Ethics [**CHRPE/AP/149/23]** and the Ghana Health Service, through the East Mamprusi Municipal Health Directorate [**GHS/NER/EMM/10/2023**]. Each participant also gave their consent to the study before data collection.

### Consent for publication

Not applicable

### Data availability

Data is available upon request to the researchers

### Authors’ contributions

Reginald A. Annan [RAA] and Charles Apprey [CA] are affiliated with the Department of Biochemistry and Biotechnology, Kwame Nkrumah University of Science and Technology, Kumasi, Ghana. Vincent Adocta Awuuh [VAA] is associated with both the Department of Biochemistry and Biotechnology at Kwame Nkrumah University of Science and Technology, Kumasi, Ghana, as a student and the Department of Nutrition and Dietetics, College of Nursing and Midwifery, Nalerigu, Ministry of Health, Ghana [P.O. Box 10, Nalerigu, Ghana]. VAA conducted the data analysis and prepared the initial manuscript draft, while RAA and CA reviewed the data analysis. All authors contributed to the study’s conceptualization, design, and interpretation of findings.

## Acknowledgements

We are deeply grateful to all study participants for their time and commitment, and we wish them continued success and well-being. We acknowledge the support of the *Canadian Queen Elizabeth II Diamond Jubilee Advanced Scholars – West Africa: Nutrition Research Capacity Building* and *Netlinks for Enhanced Health Equity and Sustainable Inclusive Growth in Rural West Africa*, whose contributions were instrumental to capacity development throughout this PhD journey. Our sincere appreciation also goes to the *Support Programme for Postgraduate Students* at North-West University, Potchefstroom, South Africa, for their role in advancing academic and research empowerment. Finally, we thank the *College of Nursing and Midwifery, Nalerigu*, for providing logistical assistance during data collection and laboratory analyses.e sincerely thank all participants in this study and wish everybody well.

## APPENDIX I

**Table 8.**
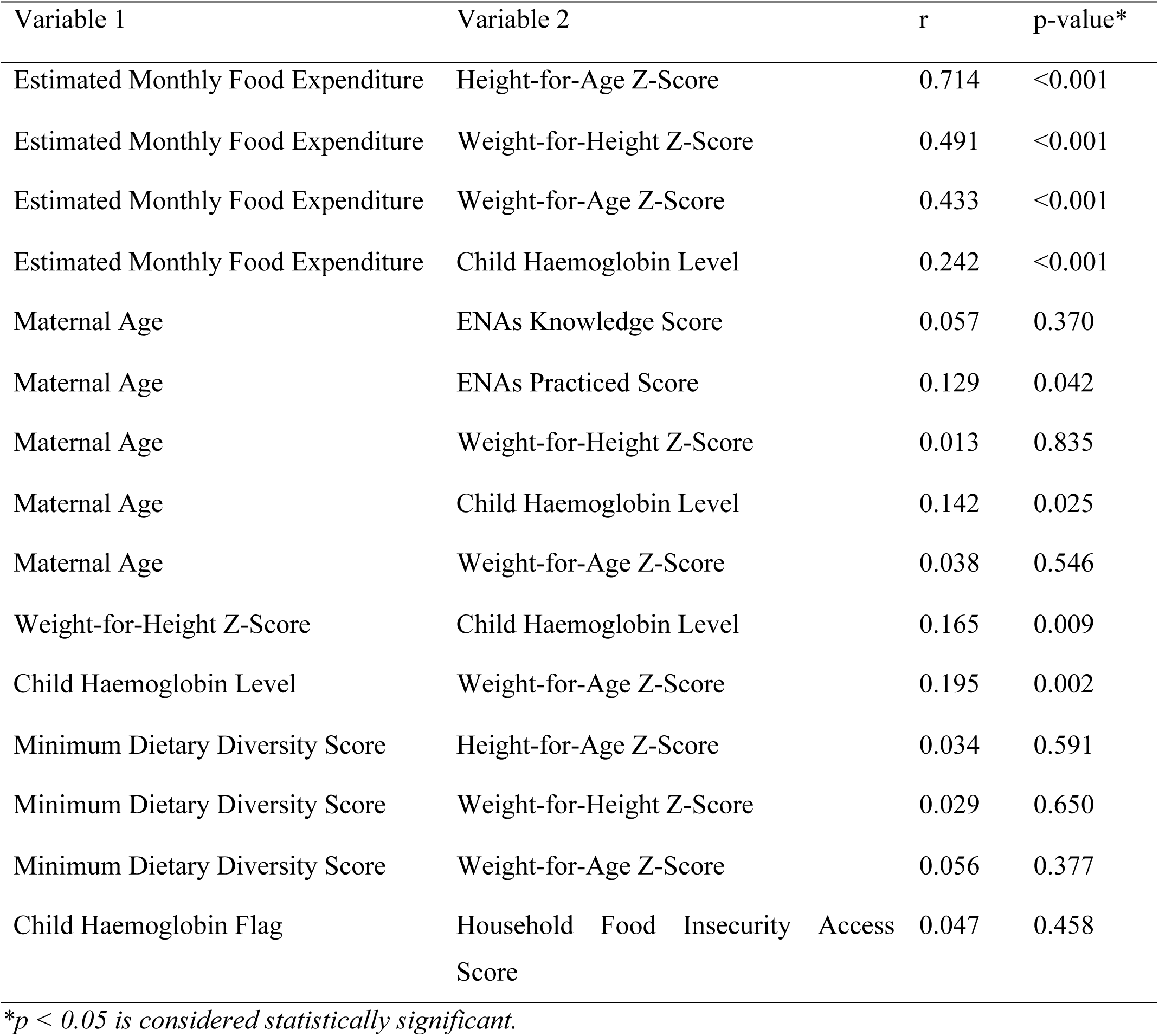
Pearson Correlation Coefficients Between Household, Maternal, and Child Nutrition Indicators.

| Variable 1 | Variable 2 | r | p-value* |
| --- | --- | --- | --- |
| Estimated Monthly Food Expenditure | Height-for-Age Z-Score | 0.714 | <0.001 |
| Estimated Monthly Food Expenditure | Weight-for-Height Z-Score | 0.491 | <0.001 |
| Estimated Monthly Food Expenditure | Weight-for-Age Z-Score | 0.433 | <0.001 |
| Estimated Monthly Food Expenditure | Child Haemoglobin Level | 0.242 | <0.001 |
| Maternal Age | ENAs Knowledge Score | 0.057 | 0.370 |
| Maternal Age | ENAs Practiced Score | 0.129 | 0.042 |
| Maternal Age | Weight-for-Height Z-Score | 0.013 | 0.835 |
| Maternal Age | Child Haemoglobin Level | 0.142 | 0.025 |
| Maternal Age | Weight-for-Age Z-Score | 0.038 | 0.546 |
| Weight-for-Height Z-Score | Child Haemoglobin Level | 0.165 | 0.009 |
| Child Haemoglobin Level | Weight-for-Age Z-Score | 0.195 | 0.002 |
| Minimum Dietary Diversity Score | Height-for-Age Z-Score | 0.034 | 0.591 |
| Minimum Dietary Diversity Score | Weight-for-Height Z-Score | 0.029 | 0.650 |
| Minimum Dietary Diversity Score | Weight-for-Age Z-Score | 0.056 | 0.377 |
| Child Haemoglobin Flag | Household Food Insecurity Access Score | 0.047 | 0.458 |

\* $p < 0.05$ is considered statistically significant.

**Table 10:** Chi-square Analysis of Factors Associated with Childhood Anaemia.

| Predictor Variable | Category | Anaemic [%] | Normal [%] | Chi-Square [$\chi^2$] | df | p-value |
| --- | --- | --- | --- | --- | --- | --- |
| Satisfaction with Maternal Health Services | Not satisfied / Satisfied | 77.4 / 57.1 | 22.6 / 42.9 | 4.634 | 1 | 0.031 |
| Quality of Maternal Health Services | Excellent / Good / Poor | 60.6 / 54.1 / 88.2 | 39.4 / 45.9 / 11.8 | 13.838 | 2 | 0.001 |
| Hb Checked at Registration | No / Yes | 78.8 / 56.7 | 21.2 / 43.3 | 5.777 | 1 | 0.016 |
| Home Support | No / Yes | 75.8 / 49.0 | 24.2 / 51.0 | 17.706 | 1 | <0.001 |
| Child Health Record Book | None / Original / Improvised | 87.5 / 57.1 / 66.7 | 12.5 / 42.9 / 33.3 | 6.031 | 2 | 0.049 |
| Household Type | Nuclear / Extended | 66.1 / 53.8 | 33.9 / 46.2 | 3.861 | 1 | 0.049 |
| Child Had Diarrhoea | Yes / No | 80.0 / 55.2 | 20.0 / 44.8 | 9.436 | 1 | 0.002 |
| Practiced Exclusive Breastfeeding | No / Yes | 79.2 / 50.9 | 20.8 / 49.1 | 17.725 | 1 | <0.001 |
| Practiced ACF | No / Yes | 68.3 / 53.5 | 31.7 / 46.5 | 5.495 | 1 | 0.019 |
| Practiced Feeding During Illness | No / Yes | 68.3 / 51.6 | 31.7 / 48.4 | 7.239 | 1 | 0.007 |
| Practiced IDA Prevention | No / Yes | 85.5 / 39.1 | 14.5 / 60.9 | 54.585 | 1 | <0.001 |
| Overall ENAs Practice | Inadequate / Adequate | 69.0 / 49.6 | 31.0 / 50.4 | 9.694 | 1 | 0.002 |
| Child Stunting Status | Stunted / Normal | 75.3 / 51.5 | 24.7 / 48.5 | 13.107 | 1 | <0.001 |
| Child Wasting Status | Wasted / Normal | 77.2 / 51.5 | 22.8 / 48.5 | 14.818 | 1 | <0.001 |
| Maternal Hb Status | Anaemic / Normal | 67.8 / 46.9 | 32.2 / 53.1 | 10.669 | 1 | 0.001 |

*Note. p < .05 indicates statistical significance. COR = Crude Odds Ratio; AOR = Adjusted Odds Ratio; ACF = Actions to Combat anaemia; IDA = Iron Deficiency Anaemia; MCV = Mean Corpuscular Volume; NEUT = Neutrophil count*

